# Nasopharyngeal microbiome composition and its clinical correlates in children hospitalized with severe pneumonia in East Africa

**DOI:** 10.1101/2025.08.27.25334445

**Authors:** Timothy O. Makori, Elijah T. Gicheru, Maureen W. Mburu, Mercy S. Sada, Omar Nyawa, Martin Mutunga, Clement Lewa, Robinson Cheruiyot, Sarah Kiguli, Peter Olupot-Olupot, Rita Muhindo, Christabel Mogaka, Thomas N Williams, Charles N. Agoti, Kathryn Maitland, Charles J. Sande

## Abstract

**Background:** Pneumonia remains the leading cause of infectious mortality in children under five, with the highest burden in sub-Saharan Africa. Dysbiosis in nasopharyngeal (NP) microbiota may influence pneumonia susceptibility and progression, but little is known about its composition or clinical relevance in low- and middle-income countries (LMICs). We characterized the NP microbiota of children hospitalized with severe pneumonia in East Africa and investigated associations with clinical outcomes.

**Methods:** We performed 16S rRNA partial gene sequencing of NP swabs collected at hospital admission from 876 children enrolled in the COAST trial across five sites in Kenya and Uganda. Clinical, demographic, and virological data were prospectively collected. Microbial profiles were analysed using hierarchical clustering, non-metric multidimensional scaling (NMDS), and multivariable regression to assess associations with respiratory viral infections, sepsis, cyanosis, bacteraemia, coma, HIV status, malnutrition, sickle cell disease (SCD), malaria, and mortality.

**Findings:** The nasopharyngeal microbiome was structured in six distinct clusters, each dominated by different genera, including *Staphylococcus*, *Streptococcus*, *Haemophilus*, *Dolosigranulum*, *Corynebacterium*, and *Moraxella*. NMDS revealed significant alignment between different microbiome clusters and key clinical outcomes: clusters dominated by *Corynebacterium* and *Dolosigranulum* were directionally associated with mortality (p < 0.001). Notably, *Corynebacterium* abundance was elevated in children who died within 48 hours of admission, then declined over longer survival intervals, approaching levels observed in survivors.

**Interpretation:** These data provide one of the largest high-resolution surveys of the paediatric upper airway microbiome in Africa and identify microbial patterns associated with viral infection, HIV status, early death and bacteraemia. The unexpected association between *Corynebacterium* and mortality may reflect distinct species composition in LMIC settings, warranting further investigation using species-resolved metagenomics. These findings lay the groundwork for microbiome-based risk stratification in childhood pneumonia.

## Introduction

Pneumonia is the leading infectious cause of death among children under five years of age, accounting for approximately 900,000 deaths globally, the majority of which occur in LMICs [1]. The mortality rate in these settings is estimated to be over 60 times higher than in high-income countries [1,2], reflecting disparities in risk factors, healthcare access, and disease management [3–5]. The aetiology of childhood pneumonia is multifactorial [6,7], with both viral and bacterial pathogens contributing to the burden of disease [7,8]. Respiratory syncytial virus (RSV) and influenza viruses are among the most frequently identified viral causes [7,9], while *Streptococcus pneumoniae* and *Haemophilus influenzae*, particularly non-vaccine types, are the most important bacterial causes of pneumonia [7,10].

The upper respiratory tract (URT) serves as the first point of contact between the host and the pathogens that cause pneumonia [11]. It is a complex ecosystem comprising both commensal and potentially pathogenic bacteria and plays a central role in respiratory health and disease by acting as a reservoir from which the lower respiratory tract may be seeded with potentially pathogenic bacteria [11–14]. Studies in high-income settings have associated respiratory illness with increased abundance of pathobionts, such as *Haemophilus influenzae* and *Streptococcus pneumoniae* [11,15–17], and reduced levels of protective taxa such as *Dolosigranulum pigrum*, *Corynebacterium spp.*, and *Moraxella lincolnii* [18–20]. However, these insights are derived almost exclusively from children in high-income countries, where demographic and social characteristics as well as environmental exposures differ substantially from children in LMICs, particularly sub-Saharan Africa [11].

In many African countries, additional risk factors such as severe malnutrition, HIV exposure, indoor air pollution, and household structure and size may exert profound effects on the respiratory microbiota and modify the risk of developing severe pneumonia [11,21]. To date, very few studies have analysed the association between the respiratory microbiome and these clinical and demographic factors within this setting. To address this gap, we conducted a comprehensive, multicentre, cross-sectional analysis of the nasopharyngeal microbiome in children hospitalized with severe pneumonia in East Africa. This study was nested within the Children’s Oxygen Administration Strategies Trial (COAST), a large randomized controlled trial evaluating oxygen delivery strategies in children with pneumonia in six hospitals in Kenya and Uganda [22]. Within this trial framework, we analysed a subset of children whose upper airways were sampled by nasopharyngeal swabbing at admission and performed bacterial community profiling using partial 16S rRNA gene sequencing. Clinical and demographic data were collected prospectively, and multiplex PCR was performed for a standardized panel of respiratory viruses. In this descriptive analysis, we sought to characterize the composition and structure of nasopharyngeal microbiota in this population and explore its associations with key clinical features, including HIV status, bacteraemia, malaria, sickle cell disease, sepsis, respiratory viral infections (RSV, Influenza and hMPV), and mortality. We provide among the first high-resolution map of the paediatric respiratory microbiome in a large-scale LMIC pneumonia cohort.

## Methods

### Study design, setting, and participants

This cross-sectional respiratory microbiome profiling study was nested within the Children’s Oxygen Administration Strategies Trial (COAST), a multicentre randomized controlled trial that evaluated different oxygen administration strategies in children with severe pneumonia [22]. The COAST trial [22] was conducted in six hospitals in East Africa: two in Kenya (Kilifi County Hospital and Coast General Teaching and Referral Hospital) and four in Uganda (Mulago National Referral Hospital, Jinja Regional Referral Hospital, Mbale Regional Referral Hospital, and Soroti Regional Referral Hospital). A subset of children aged between 28 days and 12 years who were admitted with life-threatening pneumonia and had nasopharyngeal (NP) samples collected at the point of hospital admission were selected for microbial profiling by partial 16S rRNA gene sequencing. Ethical approval was obtained from the Scientific and Ethics Review Unit of the Kenya Medical Research Institute (KEMRI) and the Uganda National Council for Science and Technology (UNCST). Written informed consent was obtained from parents and guardians in accordance with GCLP guidelines. NP samples were stored at −80°C immediately after collection for later analysis. Alongside NP sampling, detailed clinical and demographic metadata were captured, including nutritional status, vital signs, malaria status, HIV status, and outcomes such as discharge from hospital or in-hospital death. Two radiologists independently read the chest radiographs (CXRs), and a third reader resolved discrepancies and classified for presence of consolidation. A combined approach was used to define sepsis, involving diagnosis by the admitting clinician and confirmed by laboratory data of abnormal white cell counts (total WBC >12 000 or <4000 cells/µL) [23]. Sickle Cell Disease (SCD) status was confirmed by molecular genotyping. In addition, multiplex PCR testing was performed for 15 respiratory pathogens, including RSV A and B, influenza A, B, and C, parainfluenza viruses 1–4, human metapneumovirus, rhinovirus, adenovirus, coronavirus OC43 and 229E, and *Mycoplasma pneumoniae*.

### DNA extraction and 16S rRNA gene sequencing

Frozen NP swabs were thawed on ice and vortexed on a Vortex-Genie 2 mixer (Labgene Scientific). DNA was extracted using the QIAamp DNA Mini Kit (Qiagen), following mechanical lysis with glass beads on the TissueLyser II (Qiagen) and enzymatic digestion with proteinase K. DNA was eluted in AE buffer and quantified using a Qubit 4 Fluorometer (Invitrogen). Amplification of the V3–V4 hypervariable region of the bacterial 16S rRNA gene was performed using 341F and 785R primers in a 12.5 µL PCR reaction. PCR products were visualized on a 2% agarose gel, purified with AMPure XP beads (Beckman Coulter), and indexed using the Nextera XT Index Kit v2 (Illumina). Indexed libraries were purified, quantified using the Qubit dsDNA HS Assay Kit, and their size distribution assessed on the Agilent 4150 TapeStation. Libraries were normalized, pooled, and denatured with NaOH, spiked with 5% PhiX control, and sequenced on the Illumina MiSeq platform (2×300 bp, MiSeq v3 kit). Two negative controls (at extraction and PCR stages) were included for quality control.

## Data availability

The 16S rRNA sequence data generated from MiSeq sequencing in this study is available at NCBI SRA under accession number PRJNA1285267.

### Data analysis

All analyses were performed using R (v4.5.0). Raw paired-end reads were processed using the DADA2 pipeline (v1.22.0). Quality filtering and adapter trimming were performed using filterAndTriminterface of the fastqPairedFilter function, followed by error model learning with learnErrors and denoising via dada function. Forward and reverse reads were merged using mergePairs, and chimeras were removed using removeBimeraDenovo functions. Taxonomic assignment of amplicon sequence variants (ASVs) was performed using the assignTaxonomy function and the SILVA reference database (v138). The resulting ASV table was aggregated to genus level, and samples were normalized to relative abundance. Microbiome composition was assessed using hierarchical clustering of Bray–Curtis dissimilarities computed from genus-level relative abundance profiles. Clustering was performed using Ward’s method and visualised using dendrograms and stacked bar plots of the top 50 genera. Samples were assigned to six compositional clusters, identified using the NbClust R package based on the community microbiome structure. To identify dominant taxa within each cluster, average genus abundance per cluster was computed, and genus names were rendered as word clouds, scaled by mean relative abundance. To explore overall microbial structure in relation to clinical and demographic metadata, two-dimensional non-metric multidimensional scaling (NMDS) was applied to Bray–Curtis dissimilarities. The envfit function from the vegan package was used to fit clinical and virological covariates as vectors into NMDS space with 999 permutations to assess significance. Significant genus vectors were overlaid on the NMDS ordination to examine alignment of different taxa and microbial clusters with ordination gradients. Associations between individual microbial taxa and clinical or virological variable were also tested using the Maaslin2 R package (v1.21.0). Relative abundances at the genus level were used as outcomes, and different clinical features (sepsis, cyanosis, death, coma, severe malaria, HIV status, SCD, asthma history, diarrhoea, malnutrition, influenza A, RSV) were passed as fixed effects in multivariable models. Significant associations were defined using an FDR-adjusted q-value < 0.25. Results were visualised using coefficient plots (with standard errors) and annotated boxplots showing genus-level relative abundance stratified by metadata variable.

## Results

### Study population and clinical characteristics

We analysed nasopharyngeal swab (NP) samples from 876 children aged 28 days to 12 years who were admitted with life-threatening pneumonia to five hospitals in Kenya and Uganda participating in the COAST trial. The sites included Kilifi, Mulago, Jinja, Mbale, and Soroti. Baseline characteristics varied by site, with significant differences in age distribution, breastfeeding status, nutritional indicators, and mortality. Median age ranged from 7.5 months in Soroti to 14 months in Jinja. Overall mortality was highest in Kilifi and Mulago (23.1% each), and lowest in Soroti (1.7%). Severe malaria, sepsis, and coma were more frequently recorded in children from Mbale and Kilifi compared to other sites. Baseline clinical and demographic characteristics are summarized in Table 1.

**Table 1:** Baseline characteristics of the study participants.

|  | Jinja (N=78) | Kilifi (N=13) | Mbale (N=348) | Mulago (N=13) | Soroti (N=424) | P value |
| --- | --- | --- | --- | --- | --- | --- |
| **Demographic characteristics** |  |  |  |  |  |  |
| Age in months, median (IQR) | 14 (7-38.2) | 8 (3-15) | 8 (4-22) | 8 (3-15) | 7.5 (3-18) | 0.002 |
| Female, n (%) | 36 (46.2) | 4 (30.8) | 150 (43.1) | 2 (15.4) | 176 (41.5) | 0.265 |
| Breastfed, n(%) | 78 (100%) | 12 (92.3) | 347 (99.7) | 13 (100) | 343 (80.9) | <0.001 |
| Siblings, median (IQR) | 2 (1-3.8) | 4 (2-5) | 2 (1-3) | 1 (0-2) | 2 (1-4) | <0.001 |
| **Clinical characteristics** |  |  |  |  |  |  |
| Temperature, median (IQR) | 38 (37–38.5) | 37 (37–38) | 38 (37–38) | 37 (36–37) | 37 (37–38) | 0.006 |
| Heart rate, median (IQR) | 167.5 (143.8–182) | 164(158–171) | 164 (148–174) | 142 (132–158) | 157 (143–159) | <0.001 |
| Respiratory rate, median (IQR) | 60 (48–72) | 62 (58–64) | 59.5 (50.8–66) | 62 (58–66) | 63 (54–73) | 0.005 |
| Oxygen saturation, median (IQR) | 86 (78.2–88) | 89 (86–89) | 87 (81.8–89) | 83 (81–88) | 87 (83–89) | 0.105 |
| Weight, median (IQR) | 9 (7–14) | 5 (4–9) | 8 (6–10) | 8 (7–11) | 8 (6–10) | 0.048 |
| Comatose, n (%) | 3 (3.8) | 1 (7.7) | 1 (0.3) | 1 (7.7) | 2 (0.5) | <0.001 |
| Diarrhoea, n (%) | 13 (16.7) | 3 (23.1) | 69 (19.8) | 0 | 31 (7.3) | <0.001 |
| Severe malaria, n (%) | 21 (26.9) | 6 (46.2) | 87 (25) | 1 (7.7) | 24 (5.7) | <0.001 |
| Sepsis, n (%) | 16 (20.5) | 2 (15.4) | 46 (13.2) | 0 | 11 (2.6) | <0.001 |
| Cyanosis, n (%) | 2 (2.6) | 0 | 11 (3.2) | 0 | 2 (0.5) | 0.051 |
| **Comorbidities** |  |  |  |  |  |  |
| Asthma, n (%) | 0 | 1 (7.7) | 3 (0.9) | 0 | 12 (2.8) | 0.079 |
| HIV+, n (%) | 0 | 0 | 2 (0.6) | 0 | 2 (0.5) | 0.865 |
| Sickle cell crisis, n (%) | 11 (14.1) | 1 (7.7) | 15 (4.3) | 0 | 18 (4.2) | 0.005 |
| Malnutrition, muac<11.5cm, n (%) | 11 (14.1) | 5 (38.5) | 34 (9.8) | 0 | 27 (6.4) | <0.001 |
| Anaemia, n (%) | 45 (95.7) | 4 (30) | 313 (92.9) | 12 (100) | 386 (93.5) | 0.58 |
| **Outcome** |  |  |  |  |  |  |
| Died, n (%) | 5 (6.4) | 3 (23.1) | 18 (5.2) | 3 (23.1) | 7 (1.7) | <0.001 |

### Clinical associations with airway microbiome structure

To identify associations between the overall structure of the airway microbiome and clinical factors, we first conducted hierarchical clustering of genus-level relative abundance profiles and identified six distinct microbiome clusters across the 876 samples. Clustering was based on Bray–Curtis dissimilarity and Ward’s linkage method, with visualization by dendrograms and stacked bar plots of the top 50 genera as shown in Fig 1a. Six distinct clusters were identified, each displaying a unique microbial composition pattern, with one or two dominant genera typically dominating specific clusters. Word cloud visualizations (Fig 1 b) showed that cluster 1 was dominated by *Staphylococcus*, cluster 2 by *Streptococcus*, while cluster 3 was heterogeneous and exhibited no clear patterns of dominance by a single taxon. Cluster 4 was dominated by *Haemophilus*, cluster 5 by *Dolosigranulum* and *Corynebacterium*, and cluster 6 by *Moraxella* (Fig 1b). To explore the relationships between this microbial structure and host factors, we applied non-metric multidimensional scaling (NMDS) to Bray–Curtis dissimilarities of genus-level profiles. Microbiota clusters 1, 2, and 5 showed clear spatial separation in ordination space, reflecting distinct compositional patterns described above (Fig 1c). Clinical and demographic covariates were then projected into the same ordination space. Clinical outcomes including influenza A infection and death were significantly associated with ordination gradients (p < 0.05) as shown in Fig 1c. Taxonomic vectors revealed directional alignments with clinical outcomes: death was directionally associated with cluster 5 (dominated by *Corynebacterium* and *Dolosigranulum*), while influenza A infection aligned with cluster 1 (dominated by *Staphylococcus*).

**Figure 1.**
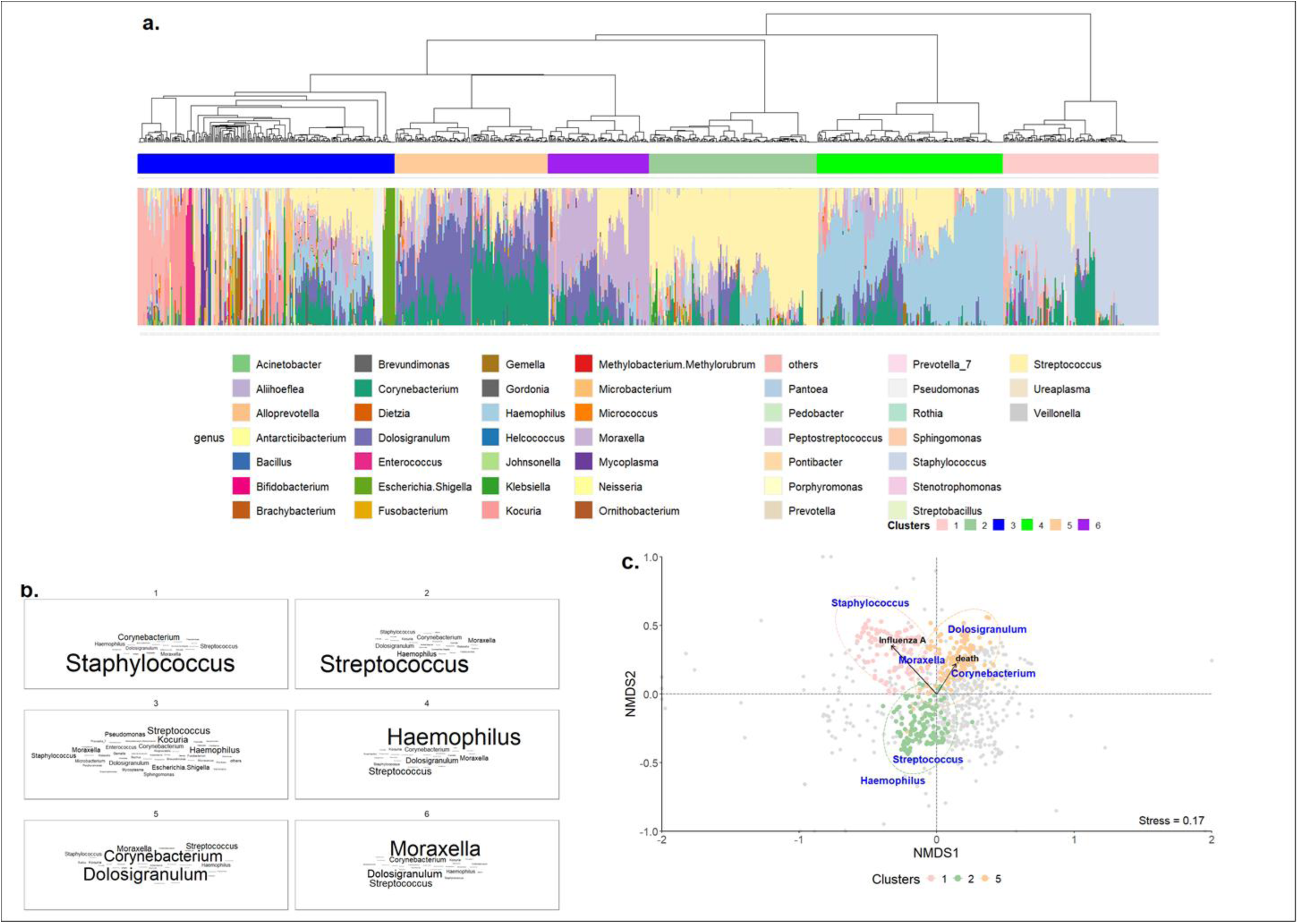
Airway microbiota structure in children hospitalized with severe pneumonia. **(a)** Hierarchical clustering of genus-level relative abundance profiles from 876 children revealed six distinct nasopharyngeal microbiome clusters based on Bray–Curtis dissimilarity and Ward’s linkage. Heatmap displays the top 50 genera; columns represent individual samples and rows represent bacterial genera. **(b)** Word clouds showing dominant genera in each cluster, scaled by mean relative abundance within cluster. Clusters were characterized by the following dominant genera: *Staphylococcus* (Cluster 1), *Streptococcus* (Cluster 2), *Haemophilus* (Cluster 4), *Corynebacterium* and *Dolosigranulum* (Cluster 5), and *Moraxella* (Cluster 6). Cluster 3 lacked a dominant genus and exhibited heterogeneous composition. **(b)** Non-metric multidimensional scaling (NMDS) of Bray–Curtis dissimilarities overlaid with microbial cluster assignments and genus vectors. Arrows represent clinical outcomes significantly associated with gradients in microbiota structure (envfit p < 0.05), including death and influenza A infection. Death vector aligned directionally with Cluster 5, while influenza A infection aligned with Cluster 1.

### Association between bacterial genera and clinical outcomes

To identify associations between specific genera and clinical outcomes, we performed multivariable regression modelling using Maaslin2 [24]. Relative abundance of genera was modelled as the outcome, with fixed effects including sepsis, cyanosis, death, coma, severe malaria, HIV status, SCD, asthma history, diarrhoea, malnutrition, RSV, and influenza A infection. Significant associations (p-value < 0.05, FDR q < 0.25) are shown in Fig 2a. *Corynebacterium* showed a positive beta association in children who died, indicating an increased abundance of this genus in the airways of children with this outcome. *Streptococcus*, on the other hand, was negatively associated with influenza A and RSV infection (Fig 2a). Similarly, *Dolosigranulum* and *Corynebacterium* were negatively associated with Influenza A infections, reflecting lower bacterial abundance in infected children, while *Gemella* showed a positive beta association among HIV positive children, indicating higher abundance in these children relative to the rest of the study population. *Haemophilus* was negatively associated with culture-confirmed bacteraemia These associations are shown using boxplots stratified by clinical group in Fig 2b.

**Figure 2.**
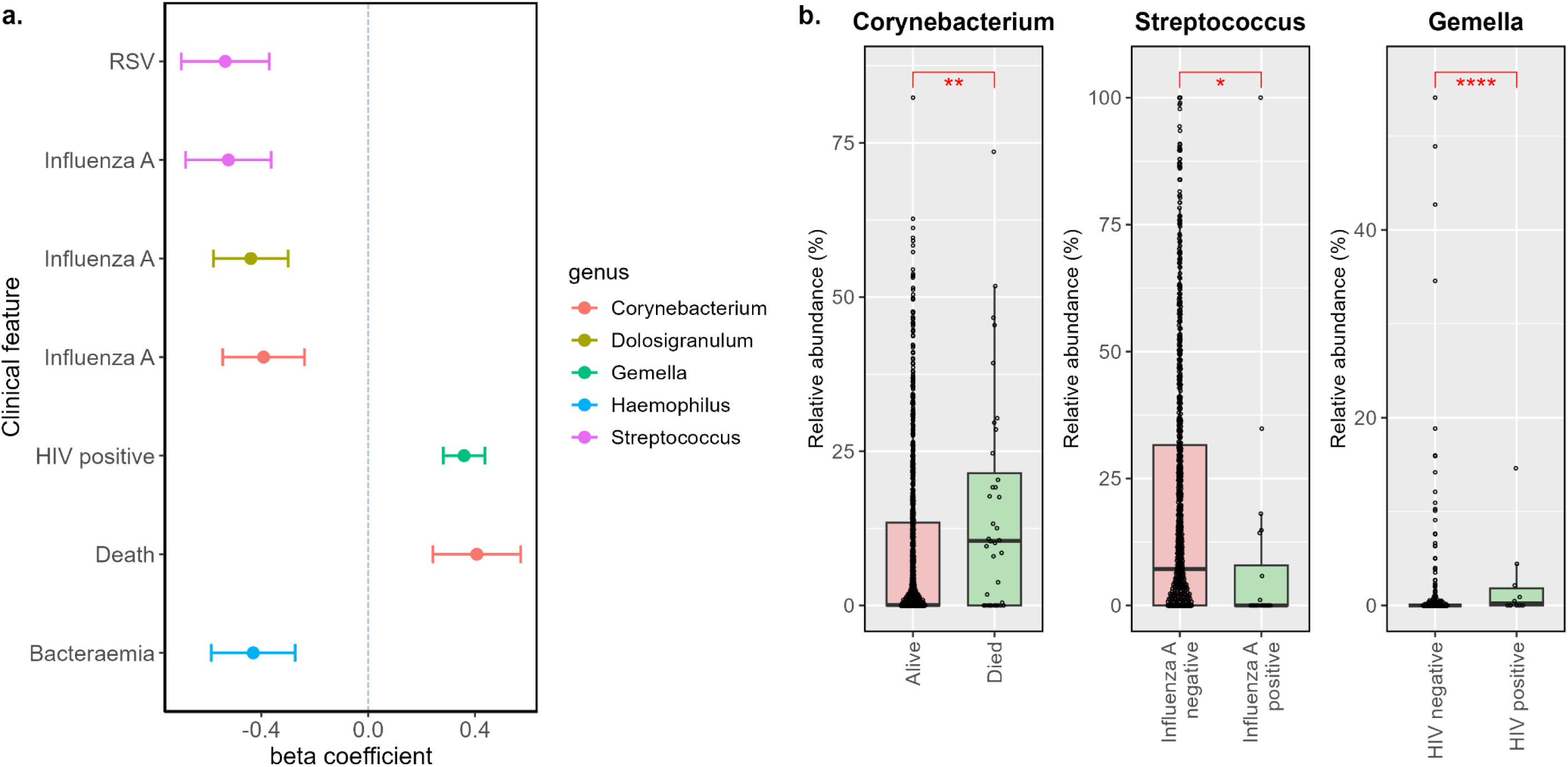
Associations between specific bacterial genera and clinical outcomes. **(a)** Results from multivariable Maaslin2 models showing associations between genus-level relative abundance and clinical or virological covariates. Beta coefficients and standard errors are shown for selected genera with a false discovery rate (FDR)–adjusted q-value < 0.25. *Corynebacterium* abundance was positively associated with death; *Gemella* was elevated in children living with HIV. *Streptococcus* was reduced in children infected with RSV and influenza A and *Haemophilus* was similarly reduced in children with culture-confirmed bacteraemia. **(b)** Annotated boxplots showing genus-level relative abundance stratified by clinical status for selected significant associations from panel (a).

### Alignment of microbial composition with time to death

To further explore the association between airway *Corynebacterium* abundance and death, we stratified children who died into five distinct time strata based on the duration of time from admission to death as follows: children who died on the day of admission, children who died one or two days after admission, children who died between 3 and 7 days after admission, and children who died between 8 and 35 days after admission. We then used nonlinear regression modelling using loess fitting to visualise changes in microbial abundance relative to the time of death. Of all the genera analysed, only *Corynebacterium* showed a clear temporal trend relative to the time of death: the highest abundance was observed among those who died on the day of admission, dropping progressively among children with longer survival times. Median *Corynebacteria* abundance in children who died ≥ 3 days after admission was similar to that of survivors (Fig 3). All other genera tested did not exhibit a clear temporal trend with time to death and were generally comparable to the median bacterial abundance of survivors (Fig 3).

**Figure 3.**
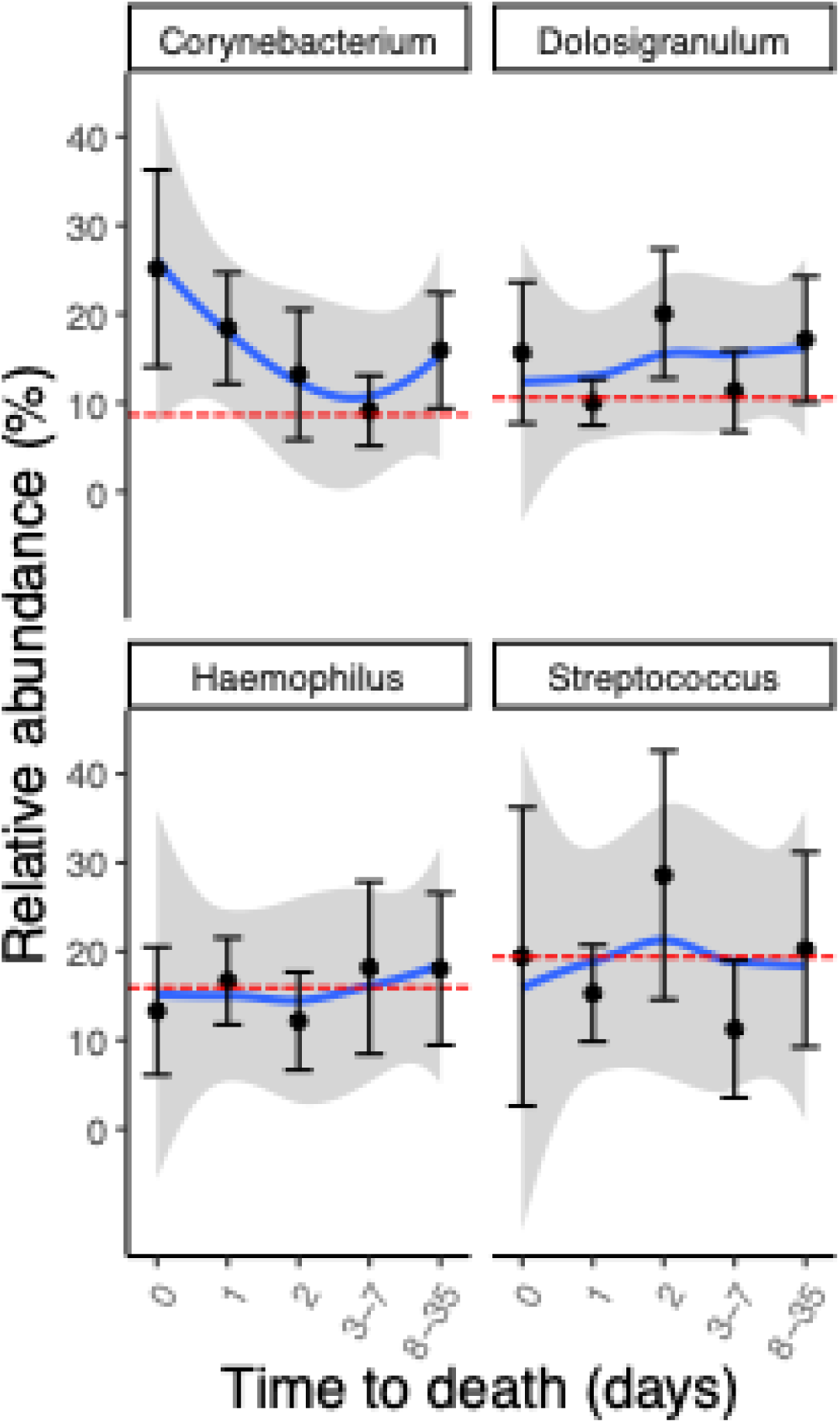
Temporal patterns of genus-level abundance by time to death. LOESS smoothed trajectories of genus-level relative abundance in children who died, stratified by time from admission to death, compared to median abundance values in survivors (red dashed lines). *Corynebacterium* abundance was highest in early deaths (day 0–2), followed by a progressive decline among later deaths, converging toward survivor levels. *Dolosigranulum*, *Haemophilus*, and *Streptococcus* abundances showed no consistent temporal variation relative to time of death. Shaded ribbons denote 95% confidence intervals for loess-smoothed estimates.

## Discussion

This study presents one of the largest high-resolution surveys of the paediatrics nasopharyngeal (NP) microbiome in sub-Saharan Africa, leveraging data from 876 children hospitalized with life-threatening pneumonia in five hospital sites in Kenya and Uganda. Using partial 16S rRNA gene sequencing, clinical phenotyping, and virological testing, we identified six microbiome clusters, characterized by varying taxonomic composition, and explored their relationships with key clinical outcomes, including sepsis, cyanosis, death, coma, severe malaria, HIV status, SCD, asthma history, diarrhoea, malnutrition, respiratory viral infections and death. This analysis showed significant associations between the airway microbiome structure and clinical outcomes, including death, HIV status, bacteraemia and influenza A infection. Genus-level associations resolved using multivariable regression modelling showed outcomes such as death and HIV status were significantly associated with different microbial taxa, including *Corynebacterium* and *Gemella*, respectively.

An unexpected finding in our study was the association between *Corynebacterium* abundance and increased mortality, particularly in children who died early (< 3 days) during hospitalization. A striking early elevation in *Corynebacterium* abundance in children who died within 48 hours, followed by a progressive decline to levels that were comparable to those of survivors among children who died later. This contrasts with the presumed beneficial role of this genus in HIC cohorts and experimental studies [25–29] and may reflect important ecological or epidemiological differences between settings. One speculative explanation is that the species composition of *Corynebacterium* differs substantially in LMICs. For example, *Corynebacterium* species such as *Corynebacterium ulcerans*, a known zoonotic pathogen [30], is more prevalent in rural African settings [31,32] due to cultural nuances that enhance human-animal interaction. *C. ulcerans* has been implicated in severe respiratory infections [33], including pneumonia [34], and produces diphtheria toxin in some cases [30,35,36]. It is typically acquired through close contact with livestock or the consumption of unpasteurized milk [36–38] - both common in rural African households where cohabitation with animals and use of raw milk as a nutritional supplement are widespread practices [39]. Recent case series from Japan and Europe have reported increasing incidence and mortality associated with *C. ulcerans* respiratory infections [34,38], underscoring its potential clinical impact. Given that our cohort included children from settings where such exposures are common, it is plausible that elevated *Corynebacterium* abundance may reflect carriage or infection with pathogenic species such as *C. ulcerans*. Unfortunately, partial 16S rRNA gene sequencing is not sufficiently sensitive for species-level resolution [40], and this hypothesis will therefore require confirmation using higher-resolution shotgun metagenomics approaches or other targeted molecular methods. We also stratified non-survivors by time to death and compared genus-specific abundance profiles among children with different survival times to those of survivors. The airway abundance of *Dolosigranulum*, *Haemophilus* and *Streptococcus*, on the other hand, did not exhibit a similar temporal pattern relative to the time of death and remained comparable to corresponding median survivor abundances at all timepoints. This temporal pattern suggests that elevated *Corynebacterium* abundance at admission may be a marker of acute disease severity, or a dysbiotic microbial state associated with imminent deterioration. In contrast, the absence of clear temporal variation in *Dolosigranulum*, *Haemophilus*, and *Streptococcus* abundance suggests that their associations with mortality are either weak, non-specific, or masked by clinical heterogeneity. Together, these findings raise the possibility that the abundance of certain *Corynebacterium* species may serve as a context-sensitive indicator of early, fulminant disease trajectories in this setting, although species-specific attributions will require confirmation using shotgun approaches as discussed above.

Several limitations of this study warrant consideration. First, this was a cross-sectional analysis at hospital admission, and therefore causality could not be inferred. Second, partial 16S rRNA sequencing does not resolve species or functional potential, limiting taxonomic specificity and precluding inference of virulence mechanisms. For instance, *Haemophilus* abundance was negatively associated with bacteraemia. While counterintuitive given the established role of *Haemophilus influenzae* as an invasive respiratory pathogen, our 16S partial sequencing approach cannot distinguish pathogenic *H. influenzae* from commensal *Haemophilus* species. Third, our data are drawn from hospitalized children with severe disease and may not generalize to mild community-acquired pneumonia or healthy controls. Nonetheless, this study provides one of the most comprehensive descriptions of the NP microbiome in African children with pneumonia to date. By combining deep clinical phenotyping with community profiling across diverse settings, we reveal both shared and context-specific features of microbial ecology in paediatric pneumonia. These findings set the stage for future mechanistic studies and highlight the importance of geographic and ecological diversity in microbiome research.

## Acknowledgments

We thank all the study participants and staff from all the centres participating in the COAST trial. We thank the Trial Steering Committee for its support of the trial.

